# Early Diagnosis and Prognosis of Cerebral Palsy From a 1-Minute Infant Video

**DOI:** 10.64898/2026.08.24.26361217

**Authors:** Colleen Peyton, Carly Luke, Arend F. Bos, Lynn Boswell, Christine Finn, Raye-Ann deRegnier, Alison Goetgeluck, Anya Gordon, Imani Mann, Kristina Stein, Megan Thorley, Roslyn N Boyd, Theresa Sukal Moulton

## Abstract

**AIM:** To evaluate whether selective motor control quantified from spontaneous infant movement recordings provides diagnostic and prognostic information for cerebral palsy (CP) beyond established movement-based assessments.

**METHOD:** This multicenter diagnostic and prognostic accuracy study included 302 infants (151 with CP) with spontaneous movement recordings obtained between 10 and 20 weeks’ corrected age from cohorts in Australia and the United States. All eligible infants with CP were included, and a comparison sample without CP was randomly selected. Recordings were scored using the Baby Observational Selective Control Appraisal (BabyOSCAR), Motor Optimality Score– Revised (MOS-R), and General Movements Assessment (GMA). Outcomes at 2 years or older included CP diagnosis, Gross Motor Function Classification System (GMFCS) level, and motor distribution.

**RESULTS:** BabyOSCAR discriminated CP diagnosis (area under the curve [AUC] 0.98), including children later classified in GMFCS level I. Among infants with CP, BabyOSCAR discriminated GMFCS levels I–II from III–V (AUC 0.89). BabyOSCAR absolute asymmetry also discriminated unilateral CP from all other infants (AUC 0.90). Diagnostic discrimination was also observed for MOS-R (AUC 0.94) and GMA (AUC 0.86).

**INTERPRETATION:** Quantifying selective motor control from brief infant movement recordings may provide complementary early information about CP diagnosis, functional level, and motor distribution.

What this paper adds:

- BabyOSCAR informs CP diagnosis, functional level, and motor distribution from brief video.
- Reduced selective motor control occurs in early infancy across diverse CP motor phenotypes.
- MOS-R provides prognostic information about later gross motor function and distribution.

---

Early identification of cerebral palsy (CP) enables timely access to intervention and family-centered care during a critical period of neurodevelopment. Advances in early detection have led to the development of clinical pathways that incorporate neuroimaging, neurological examination, and observation of spontaneous movement, with the General Movements Assessment (GMA) demonstrating high accuracy for identifying CP in early infancy^1,2^.

While GMA is highly effective at identifying CP in infants^1,2^, it provides a categorical assessment of movement quality and more limited information regarding later functional level or distribution of motor involvement. Detailed early prognostic information about these outcomes may support treatment planning and family counseling.

Detailed approaches to evaluating spontaneous movement, such as the Motor Optimality Score–Revised (MOS-R)^3^, offer additional granularity by characterizing movement patterns and identifying features associated with CP. These approaches, however, may be limited in their ability to capture specific aspects of motor control that are directly related to underlying motor pathway dysfunction and later functional outcomes.

Selective motor control (SMC), defined as the ability to move joints independently, is a key determinant of functional level in CP^4^ and reflects the integrity of descending motor pathways^5,6^. Early manifestations of SMC can be observed in spontaneous infant movement^7,8^, providing an opportunity to quantify emerging motor control during a critical developmental window. The Baby Observational Selective Control AppRaisal (BabyOSCAR) is a standardized observational tool designed to measure independent joint motion from brief video recordings (one minute) of spontaneous movement in early infancy^7^.

Prior work has demonstrated that BabyOSCAR scores at 3 months corrected age are predictive of CP diagnosis, functional level, and body distribution at 2 years^9^. These findings, however, were established in a smaller cohort, restricted to infants with spastic CP^9^.

The aim of the present study was to evaluate the diagnostic and prognostic performance of BabyOSCAR in a larger, multicenter cohort of infants with and without CP, including a broader range of CP motor subtypes. We also compared BabyOSCAR with established tools derived from the same spontaneous movement recordings, including GMA and MOS-R, to evaluate their discrimination of CP diagnosis, functional level, and motor distribution.

## Method

### Study design and participants

This multicenter diagnostic and prognostic accuracy study included infants with spontaneous movement recordings obtained between 10 and 20 weeks’ corrected age from research cohorts at The University of Queensland, Australia, and Northwestern University, United States, between March 2019 and March 2024. None of the infants were included in the previous BabyOSCAR diagnostic and prognostic accuracy study^9^. All eligible infants with confirmed CP at follow-up at 2 years or older were included. From eligible infants without CP, an equal-sized comparison sample was randomly selected. Infants were eligible if a spontaneous movement recording of sufficient quality for scoring and follow-up outcome data at 2 years or older were available. Infants were excluded if they had joint contractures or peripheral nerve injuries limiting joint motion due to non-central nervous system causes.

CP diagnosis, Gross Motor Function Classification System (GMFCS) level, and motor distribution (unilateral or bilateral) were determined at follow-up. In Australia, outcomes were determined by trained clinicians using Australian Cerebral Palsy Register criteria^10^. In the United States, CP diagnosis and motor distribution were determined by physicians through clinical examination; GMFCS level was determined by trained clinicians using clinical assessment and parent report. These outcome assessments preceded development of the BabyOSCAR; therefore, assessors were blinded to BabyOSCAR scores.

Institutional review board approval was obtained at all participating sites, with written informed consent obtained or waived as approved.

### Measures

#### BabyOSCAR

BabyOSCAR (Observational Selective Control AppRaisal) is a standardized observational assessment of the capacity for independent joint motion during spontaneous movement. Independent joint motion is defined as movement at an individual joint without concurrent movement at other joints within the same limb and without mirroring in the contralateral joint^7^.

Independent movement at each joint on the left and right sides is scored as present (1) or absent (0), yielding a total score ranging from 0 to 32, with higher scores indicating greater capacity for independent joint motion. An asymmetry score is calculated as the difference between the number of joints demonstrating independent motion on the left and right sides^9^. An absolute value of the asymmetry score was used for analysis.

### General Movement Assessment and Motor Optimality Score-Revised

The General Movement Assessment (GMA) classifies fidgety movements between 10 and 20 weeks corrected age as normal, sporadic, abnormal or absent^11,12^. For the prespecified binary analysis, fidgety movements were classified as normal or aberrant fidgety.

The Motor Optimality Score–Revised (MOS-R), provides a detailed assessment of spontaneous motor behavior between 10 and 20 weeks corrected age^3^. In addition to fidgety movements, the MOS-R evaluates observed movement patterns, age-adequate motor repertoire, observed postural patterns, and movement character. Domain scores are summed to yield a total score from 5 to 28, with higher scores indicating more optimal motor performance^13^.

Two MOS-R items, Segmental Movements of Fingers and Wrists and Asymmetry of Finger Posture, were defined a priori as asymmetry features, and their combined presence was used for asymmetry analysis.

### Video scoring procedures

BabyOSCAR, GMA, and MOS-R were all scored from the same video recordings. BabyOSCAR was scored from a 1-minute segment during which all limbs were visible. GMA and MOS-R were scored from approximately 3–5 minutes recordings, consistent with standard assessment procedures^11^. Infants were filmed supine and wearing clothing that allowed visualization of hands and feet. Raters were masked to infant medical history and outcome. All assessments were scored by trained raters who had met reliability requirements before study scoring.

### Statistical Analysis

All analyses were conducted in R (version 2022.07.2). Continuous variables were summarized using medians and interquartile ranges and categorical variables using counts and percentages. Group differences were assessed using Wilcoxon rank-sum tests for continuous variables and χ² or Fisher exact tests for categorical variables, as appropriate.

Diagnostic and prognostic discrimination of BabyOSCAR, MOS-R, and GMA was evaluated using receiver operating characteristic (ROC) curve analyses. Areas under the curve (AUCs) with 95% confidence intervals (CIs) were calculated, and differences between tools were compared using DeLong tests. Data-derived thresholds were identified using the Youden index. Sensitivity, specificity, positive predictive value, and negative predictive value were reported with 95% CIs. To evaluate performance when minimizing false-positive classifications, sensitivity was additionally estimated at a fixed specificity of 90%, with 95% CIs estimated using 2000 stratified bootstrap replicates.

Analyses evaluated (1) discrimination of CP diagnosis in the full sample; (2) discrimination of Gross Motor Function Classification System (GMFCS) levels I–II versus III–V among infants with CP; and (3) discrimination of unilateral CP from all other infants using asymmetry measures.

Associations between asymmetry measures and unilateral CP were evaluated using logistic regression, with BabyOSCAR absolute asymmetry score and the prespecified combined MOS-R asymmetry variable examined in separate and combined models.

A post hoc exploratory analysis examined infants whose early BabyOSCAR scores suggested a different functional classification from their observed GMFCS level at follow-up. Clinical histories were reviewed descriptively to identify factors or concurrent diagnoses potentially associated with discordant findings. Reporting followed the STARD guidelines for diagnostic accuracy studies.

## Results

### Participants

A total of 302 infants were included, 151 with CP and 151 without CP. Gestational age, birthweight, and sex did not differ between groups. Infants with CP were assessed at a slightly older postmenstrual age than infants without CP (Table 1).

**Table 1.** Characteristics among 302 infants with and without cerebral palsy.

|  | Overall<br>N=302 | No CP<br>N=151 | CP<br>N= 151 | p-value |
| --- | --- | --- | --- | --- |
| Median Gestational age, weeks (IQR) | 34 (28, 38) | 34 (29, 38) | 33 (27, 38) | 0.2<sup>a</sup> |
| Median Birthweight, g (IQR) | 2187 (1086, 3134) | 2370 (1157, 3190) | 2101 (1045, 3090) | 0.3<sup>a</sup> |
| Sex, Male (%) | 176 (58) | 91 (60) | 85 (56) | 0.6<sup>b</sup> |
| Median postmenstrual age at testing, weeks (IQR) | 54 (53, 54) | 54 (52, 54) | 54 (53, 55) | 0.005<sup>a</sup> |
| Median BabyOSCAR total score (IQR) | 22 (14, 26) | 26 (24, 32) | 15 (9, 19) | <0.001<sup>a</sup> |
| Median MOS-R score (IQR) | 20 (7, 24) | 24 (22, 25) | 7 (6, 10) | <0.001<sup>a</sup> |

CP, cerebral palsy; IQR, interquartile range; MOS-R, Motor Optimality Score–Revised.
□ Wilcoxon rank-sum test.
□ $\chi^2$ test.

Among infants with CP, GMFCS distribution was level I (42%), II (10%), III (14%), IV (16%), and V (19%). Characteristics by GMFCS group are presented in Table 2. Gestational age, birthweight, and sex did not differ between infants classified in GMFCS levels I–II and III–V. Infants in GMFCS levels I–II were more likely to have unilateral and spastic CP, whereas those in levels III–V more frequently had bilateral involvement and dyskinetic or mixed motor types (Table 2). Among infants with aberrant fidgety movements (n=126), 89 (70%) had absent, 36 (29%) sporadic, and one (1%) abnormal fidgety movements.

**Table 2.** Characteristics among 151 infants with cerebral palsy GMFCS levels I-II and III-V.

|  | Overall<br>N=151 | GMFCS I-II<br>N=78 | GMFCS III-V<br>N=73 | p-value |
| --- | --- | --- | --- | --- |
| Median gestational age, weeks (IQR) | 33 (27, 38) | 31 (26, 38) | 35 (28, 38) | 0.06<sup>a</sup> |
| Median birthweight, g (IQR) | 2101 (1045, 3090) | 1795 (924, 2798) | 2210 (1100, 3245) | 0.2<sup>a</sup> |
| Sex, Male (%) | 85 (56) | 41 (53) | 44 (60) | 0.4<sup>b</sup> |
| Median Postmenstrual age at testing, weeks (IQR) | 54 (53, 55) | 54 (53, 55) | 54 (53, 55) | 0.5<sup>a</sup> |
| Median BabyOSCAR total score (IQR) | 15 (9, 19) | 18 (15, 20) | 9 (4, 13) | <0.001<sup>a</sup> |
| Median MOS-R score (IQR) | 7 (6, 10) | 10 (7, 21) | 6 (6, 7) | <0.001<sup>a</sup> |
| CP distribution, Unilateral (%) | 60 (40) | 52 (67) | 8 (11) | <0.001<sup>b</sup> |
| Motor type, n (%) |  |  |  | 0.02<sup>c</sup> |
| Spastic | 73 (49) | 45 (58) | 28 (39) |  |
| Dyskinetic | 32 (21) | 15 (19) | 17 (24) |  |
| Mixed | 35 (23) | 11 (14) | 24 (33) |  |
| Hypotonic | 2 (1) | 1 (1) | 1 (1) |  |
| Unknown | 8 (5) | 6 (8) | 2 (3) |  |

CP, cerebral palsy; GMFCS, Gross Motor Function Classification System; IQR, interquartile range; MOS-R, Motor Optimality Score-Revised.
□ Wilcoxon rank-sum test
□ $\chi^2$ test
□ Fisher exact test

### Diagnostic discrimination of CP

BabyOSCAR, MOS-R, and GMA fidgety classification discriminated CP from no CP. BabyOSCAR had an AUC of 0.98 (95% CI 0.97–1.00), compared with 0.94 (95% CI 0.91–0.96) for MOS-R and 0.86 (95% CI 0.83–0.90) for GMA. BabyOSCAR demonstrated greater discrimination than MOS-R (difference in AUC, 0.05; 95% CI 0.02–0.07; p<0.001) and GMA (difference in AUC, 0.12; 95% CI 0.08–0.16; p<0.001); MOS-R also demonstrated greater discrimination than GMA (difference in AUC, 0.07; 95% CI 0.05–0.10; p<0.001; Figure 1).

**Figure 1.**
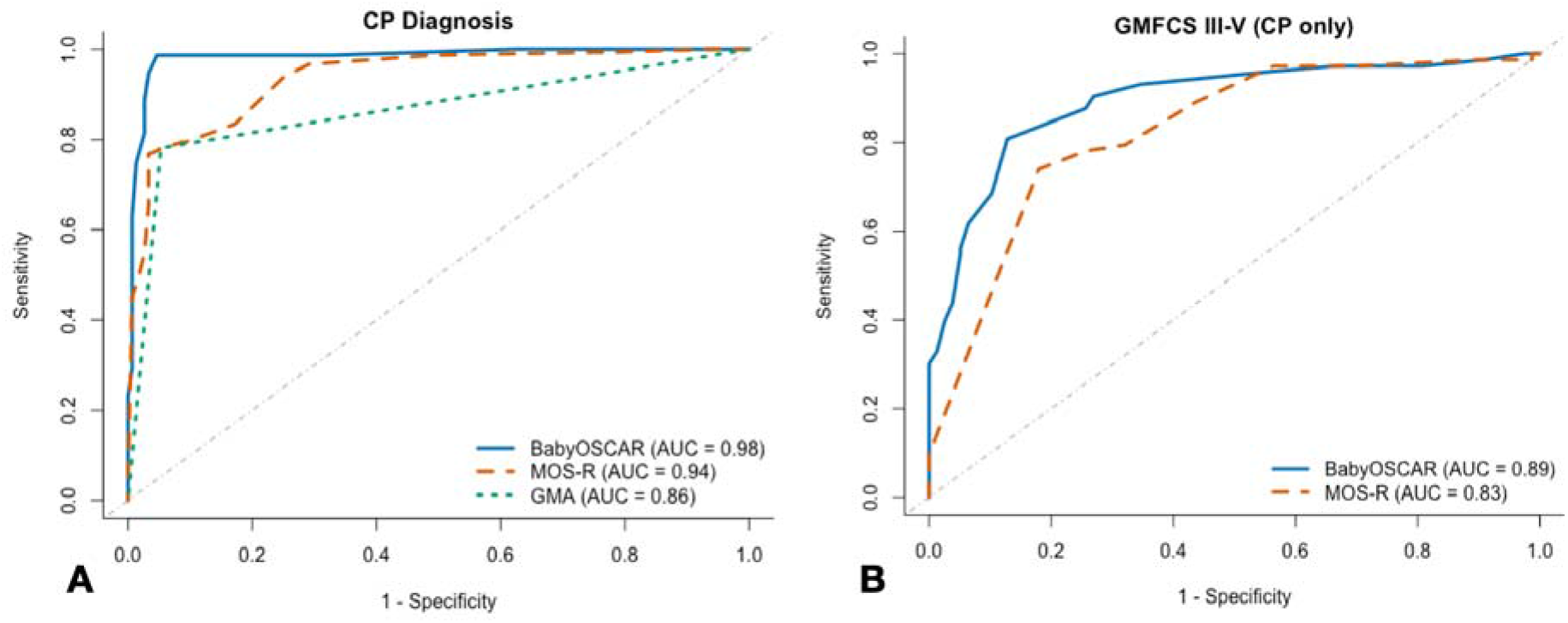
ROC Curves for Cerebral Palsy Diagnosis and GMFCS Level Using General Movements Assessment, BabyOSCAR, and MOS-R. **(A)** Receiver Operating Characteristic (ROC) curves for identification of cerebral palsy using General Movements Assessment (fidgety), BabyOSCAR, and MOS-R (n=302) **(B)** ROC curves for discrimination of GMFCS level I–II versus III–V among infants with cerebral palsy using BabyOSCAR and MOS-R (n=151).

At the data-derived Youden thresholds, BabyOSCAR had higher sensitivity than MOS-R and GMA with comparable specificity (Table 3). At 90% specificity, BabyOSCAR maintained higher sensitivity than MOS-R (Table 3).

**Table 3.**
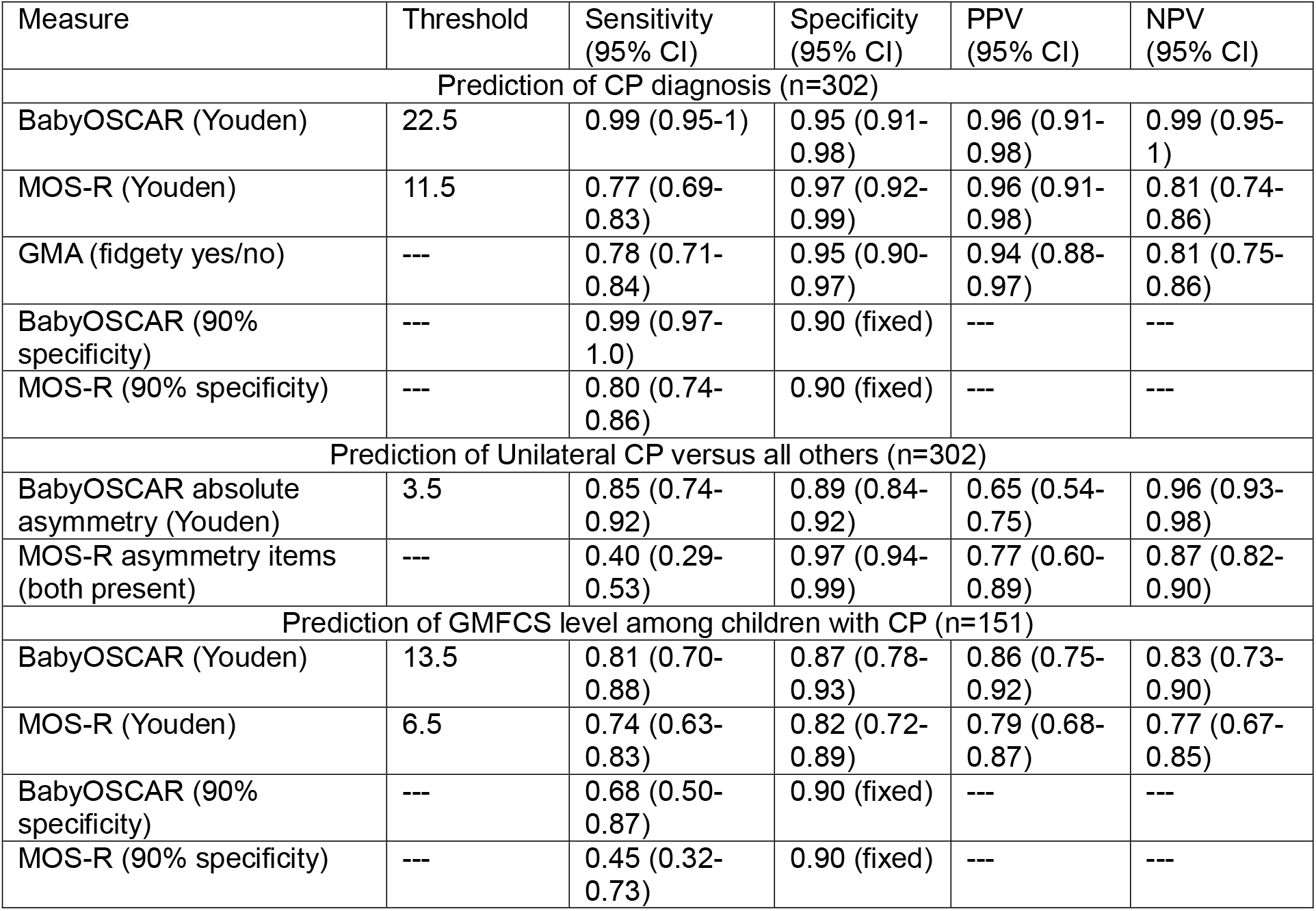
Diagnostic Accuracy for Cerebral Palsy and Prognostic Accuracy for Functional Level and Motor Distribution.

| Measure | Threshold | Sensitivity (95% CI) | Specificity (95% CI) | PPV (95% CI) | NPV (95% CI) |
| --- | --- | --- | --- | --- | --- |
| Prediction of CP diagnosis (n=302) |  |  |  |  |  |
| BabyOSCAR (Youden) | 22.5 | 0.99 (0.95-1) | 0.95 (0.91-0.98) | 0.96 (0.91-0.98) | 0.99 (0.95-1) |
| MOS-R (Youden) | 11.5 | 0.77 (0.69-0.83) | 0.97 (0.92-0.99) | 0.96 (0.91-0.98) | 0.81 (0.74-0.86) |
| GMA (fidgety yes/no) | --- | 0.78 (0.71-0.84) | 0.95 (0.90-0.97) | 0.94 (0.88-0.97) | 0.81 (0.75-0.86) |
| BabyOSCAR (90% specificity) | --- | 0.99 (0.97-1.0) | 0.90 (fixed) | --- | --- |
| MOS-R (90% specificity) | --- | 0.80 (0.74-0.86) | 0.90 (fixed) | --- | --- |
| Prediction of Unilateral CP versus all others (n=302) |  |  |  |  |  |
| BabyOSCAR absolute asymmetry (Youden) | 3.5 | 0.85 (0.74-0.92) | 0.89 (0.84-0.92) | 0.65 (0.54-0.75) | 0.96 (0.93-0.98) |
| MOS-R asymmetry items (both present) | --- | 0.40 (0.29-0.53) | 0.97 (0.94-0.99) | 0.77 (0.60-0.89) | 0.87 (0.82-0.90) |
| Prediction of GMFCS level among children with CP (n=151) |  |  |  |  |  |
| BabyOSCAR (Youden) | 13.5 | 0.81 (0.70-0.88) | 0.87 (0.78-0.93) | 0.86 (0.75-0.92) | 0.83 (0.73-0.90) |
| MOS-R (Youden) | 6.5 | 0.74 (0.63-0.83) | 0.82 (0.72-0.89) | 0.79 (0.68-0.87) | 0.77 (0.67-0.85) |
| BabyOSCAR (90% specificity) | --- | 0.68 (0.50-0.87) | 0.90 (fixed) | --- | --- |
| MOS-R (90% specificity) | --- | 0.45 (0.32-0.73) | 0.90 (fixed) | --- | --- |

### Prognostic discrimination of GMFCS level

Among infants with CP, both measures discriminated between GMFCS levels I–II and III–V. BabyOSCAR demonstrated greater discrimination than MOS-R (AUC, 0.89 [95% CI, 0.84–0.95] vs 0.83 [95% CI, 0.76–0.89]; difference in AUC, 0.07 [95% CI, 0.001–0.13]; p=0.046).

At the data-derived Youden thresholds, BabyOSCAR had higher sensitivity and specificity than MOS-R, with greater differences when specificity was constrained to 90% (Table 3).

### Adjustment for postmenstrual age

After adjustment for postmenstrual age at testing, BabyOSCAR remained associated with CP diagnosis (OR 0.42, 95% CI 0.32–0.51; *p*<0.001), while postmenstrual age was not (*p*=0.43). For MOS-R, score remained associated with CP diagnosis (OR 0.74, 95% CI 0.69–0.79; *p*<0.001), while the association with postmenstrual age did not reach statistical significance (*p*=0.06). Model discrimination was unchanged after adjustment for postmenstrual age for BabyOSCAR (AUC 0.98) and MOS-R (AUC 0.94).

### Prognostic discrimination of motor distribution

BabyOSCAR absolute asymmetry scores were higher among infants with unilateral CP than among all other infants (median 6 [IQR 4–8.25] vs 1 [IQR 1–2]). Both prespecified MOS-R asymmetry features were present in 24 of 60 (40%) infants with unilateral CP compared with 7 of 242 (3%) other infants. Higher BabyOSCAR absolute asymmetry scores were associated with unilateral CP (OR 2.17 per 1-point increase, 95% CI 1.82–2.67; p<0.001). Prespecified MOS-R asymmetry features were also associated with unilateral CP in univariable analysis (OR 22.38, 95% CI 9.42–59.78; p<0.001). BabyOSCAR absolute asymmetry demonstrated greater discrimination of unilateral CP from all other infants than MOS-R asymmetry features (AUC 0.90, 95% CI 0.85–0.95 vs 0.69, 95% CI 0.62–0.75; difference in AUC 0.22, 95% CI 0.15–0.29; p<0.001; Figure 2).

**Figure 2.**
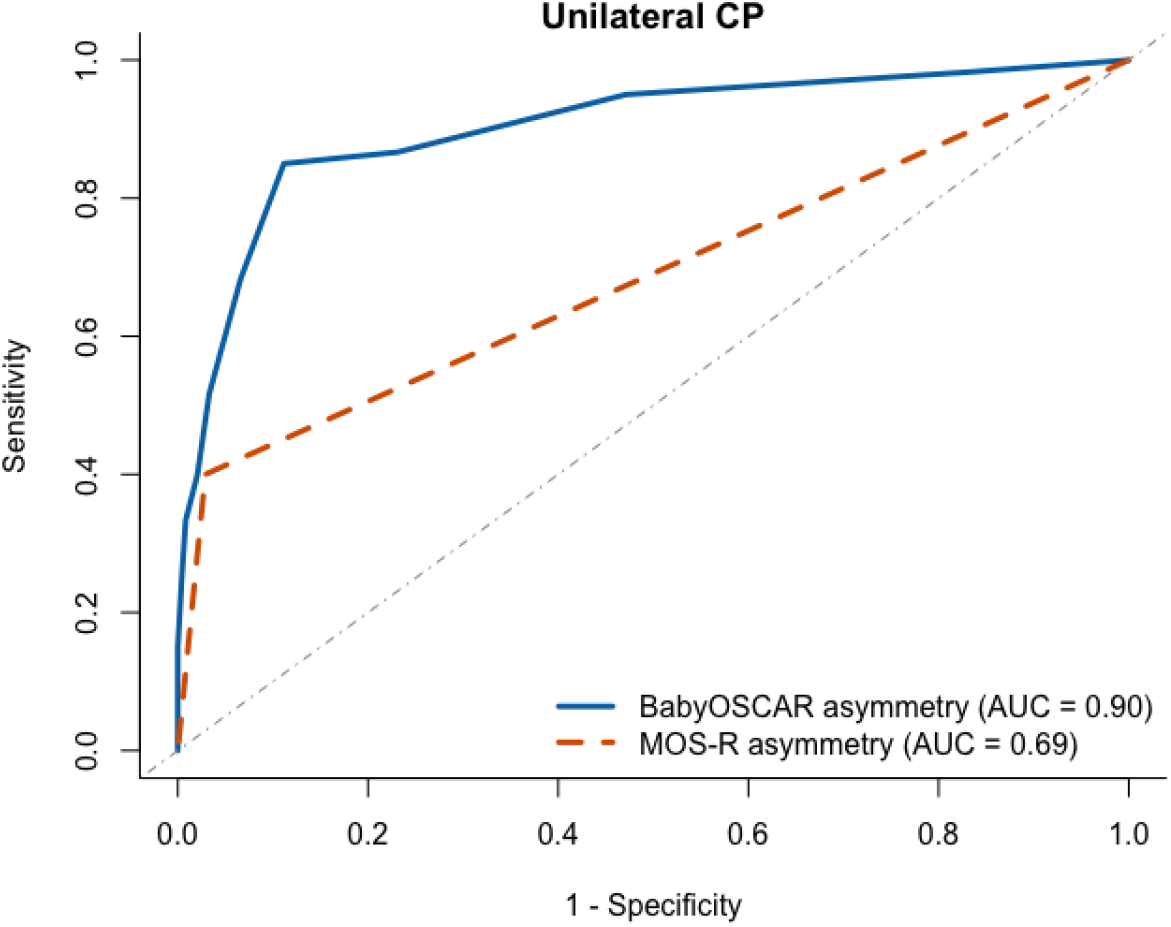
Comparative ROC Curves for BabyOSCAR and MOS-R in Identifying Unilateral Cerebral Palsy. Receiver Operating Characteristic (ROC) curves for identification of unilateral cerebral palsy using BabyOSCAR absolute asymmetry score, and MOS-R combined asymmetry features (n=302).

In the combined model, BabyOSCAR absolute asymmetry remained independently associated with unilateral CP (OR 2.07, 95% CI 1.72–2.56; p<0.001), whereas MOS-R asymmetry features were not (OR 3.39, 95% CI 0.98–12.03; p=0.054).

The data-derived BabyOSCAR absolute asymmetry threshold was 3.5 (corresponding to a score ≥4), yielding 85% sensitivity and 89% specificity. In contrast, MOS-R asymmetry features demonstrated lower sensitivity (40%) but higher specificity (97%; Table 3).

### Exploratory review of GMFCS discordance

Among infants whose BabyOSCAR scores suggested GMFCS levels I–II but who were later classified in GMFCS levels III–V (n=14), eight had a history of neonatal seizures, six had dyskinetic or mixed CP, and four had genetic syndromes and/or congenital anomalies.

Among infants whose BabyOSCAR scores suggested GMFCS levels III–V but who were later independently ambulant (n=10), six had dyskinetic or mixed motor features and two had a history of seizures. Of the 24 infants misclassified by BabyOSCAR, 12 (50%) were also misclassified by MOS-R. MOS-R misclassified 33 infants overall.

## Discussion

In this multicenter diagnostic and prognostic accuracy study, quantification of selective motor control from a brief infant video demonstrated high discrimination of CP diagnosis, functional level, and motor distribution. BabyOSCAR demonstrated greater diagnostic discrimination than GMA and MOS-R and maintained high sensitivity and specificity. Importantly, these findings extend prior work^9^ by demonstrating that BabyOSCAR diagnostic and prognostic performance remains robust in an independent cohort that more than doubled the sample size and included spastic, dyskinetic, and mixed CP motor types, with data-derived cut points and performance similar to those identified in our initial study^9^.

Differences in diagnostic performance compared with prior studies likely reflect differences in cohort composition. Previous work has reported very high sensitivity and specificity for MOS-R in detecting CP^14^, although these findings were derived from samples with relatively few confirmed CP cases. The present cohort included a substantial proportion of children later classified as GMFCS I. Consistent with this different case mix, GMA sensitivity in the current study (0.78) was lower than that reported in many prior studies (>90%)^2^, which may reflect the greater representation of children with GMFCS I. Prior work has shown lower GMA sensitivity in this group^3,15^. Together, these findings highlight the importance of case mix, particularly the distribution of functional levels, when comparing diagnostic performance across studies. In this context, the maintained performance of BabyOSCAR may reflect its ability to capture early differences in selective motor control that remain detectable even among children later classified as GMFCS I, whose early motor differences may be more subtle.

Among infants with CP, both BabyOSCAR and MOS-R discriminated later functional level, with greater discrimination for BabyOSCAR in distinguishing GMFCS I–II from III–V. BabyOSCAR performance for functional classification was lower than previously reported in our cohort restricted to infants with spastic CP^9^. Exploratory analyses suggest that discordance between early BabyOSCAR scores and later GMFCS classification occurred more frequently among infants with dyskinetic or mixed type CP and those with a history of neonatal seizures. In particular, neonatal seizures were common among infants whose early BabyOSCAR scores suggested independent ambulation but who were later classified with GMFCS III-V, raising the possibility that evolving neurologic factors contribute to differences between early motor performance and later functional outcomes^16,17^. Although these findings require further study, they reinforce the importance of considering the broader neurological and clinical context when using early motor assessments to inform prognostic conversations with families.

The present study also extends prior work by evaluating asymmetry as a marker of CP distribution. BabyOSCAR absolute asymmetry strongly discriminated unilateral CP from all other infants and demonstrated greater discriminative performance than MOS-R asymmetry features. The BabyOSCAR asymmetry cut point was lower than previously reported^9^, with high sensitivity and specificity in this broader cohort. In contrast, MOS-R asymmetry features demonstrated high specificity but limited sensitivity, suggesting that while these features may identify more pronounced asymmetry, they may miss subtler but clinically relevant differences. This may reflect differences in the scope of measurement, as MOS-R asymmetry features are derived primarily from distal upper extremity postures, whereas BabyOSCAR evaluates asymmetry across multiple joints throughout the upper and lower extremities using a continuous measure.

These findings likely reflect the construct each tool captures. GMA and MOS-R characterize spontaneous movement quality, whereas BabyOSCAR quantifies independent joint control, a feature more closely aligned with descending motor pathway dysfunction in CP. Prior work suggests that isolated joint motion reflects corticospinal integrity^5,6^ and is strongly related to later function^9,8,4^, which may explain the greater precision of BabyOSCAR for diagnosis, GMFCS level, and distribution. Notably, GMA, MOS-R and BabyOSCAR can all be derived from the same spontaneous movement recording, allowing for the extraction of multiple clinically relevant features from a single video. From a clinical perspective, GMA remains a rapid and effective screening tool, whereas BabyOSCAR may provide additional precision for prognosis, functional classification, and distribution, particularly when CP is identified or suspected. Together, these findings support a complementary approach in which multiple aspects of early motor function can be efficiently evaluated from the same scalable, video-based assessment.

Importantly, the present cohort included children with dyskinetic and mixed motor types, extending evaluation of selective motor control beyond predominantly spastic CP. In our prior prospective longitudinal study, reduced selective motor control was also observed in two infants who later developed dyskinetic or mixed CP (one each)^8^, although this small number precluded conclusions regarding motor type. Together, these observations suggest that while selective motor control is closely linked to corticospinal system integrity, reduced selective motor control in early infancy may also occur in the context of broader motor network dysfunction across different CP motor phenotypes.

Future studies integrating longitudinal measures of selective motor control with neuroimaging and established clinical assessments such as the HINE^18^ may help clarify how different patterns of brain injury contribute to these early motor phenotypes and whether these measures provide complementary prognostic information. The use of brief video recordings also creates opportunities for remote implementation. Prior work using the Baby Moves app has demonstrated the feasibility of obtaining parent-recorded infant movement videos in the home^19^, supporting the potential for similar approaches to be evaluated with BabyOSCAR. Videos could be evaluated remotely by trained clinicians, while automated video analysis and machine-learning approaches may further support scalable quantification of selective motor control, particularly where specialized expertise is not locally available. Prospective validation in unselected and population-based cohorts will be important to determine performance in real-world early detection pathways.

This study has several strengths, including a multicenter design, evaluation in a cohort independent of the initial BabyOSCAR diagnostic and prognostic study, use of standardized video recordings scored across multiple tools, masked outcome ascertainment, and inclusion of children across CP motor types and functional levels. Several limitations should also be considered. The study included equal numbers of infants with and without CP; therefore, positive and negative predictive values reflect the prevalence of CP in this study sample and should not be generalized to populations with different CP prevalence. Although the similarity of BabyOSCAR cut points and performance across independent cohorts is encouraging, these thresholds require prospective validation in unselected clinical populations. Finally, the exploratory analyses of discordant functional classifications involved small subgroups and should be considered hypothesis-generating.

### Conclusions and Relevance

Quantifying selective motor control from spontaneous movement recordings in early infancy provided strong discrimination of CP diagnosis, functional level, and motor distribution across diverse CP motor types. Used alongside GMA, BabyOSCAR may provide additional prognostic precision to support individualized early intervention planning and family counselling within scalable, video-based early detection pathways.

## Data Availability

All data produced in the present study are available upon reasonable request to the authors

